# Cardiac Arrhythmia Associated with Psychoactive Drugs: An analysis of the FDA Adverse Event Reporting System

**DOI:** 10.64898/2026.09.08.26362577

**Authors:** Mori J Krantz, Mark C Haigney, Mary Ross Southworth, Norman Stockbridge, David P Kao

## Abstract

**Background:** Several Schedule I substances have demonstrated potential benefits for refractory psychiatric conditions and addiction. Some have been legalized at the state level, while others like kratom (mitragynine) are available over-the-counter in some states and illegal in others. A recent presidential executive order directs certain federal agencies to accelerate development and facilitate access to psychedelic drugs through increased funding and expedited FDA review. Studies investigating the therapeutic potential of these drugs are limited and focused disproportionately on efficacy over safety. Rigorous evaluation of cardiovascular safety, including potential arrhythmia liability, is essential.

**Methods:** We analyzed reports from the FDA Adverse Event Reporting System from 2000-2024. involving Schedule 1 and other unscheduled drugs with purported therapeutic potential including 3,4-methylenedioxy-methamphetamine (MDMA), mitragynine, ibogaine, lysergic acid diethylamide (LSD), mescaline, psilocybin, tetrahydrocannabinol (THC), and dimethyltryptamine (DMT). We used FDA-approved drugs (e.g., dofetilide, naltrexone) as quasi-experimental positive and negative controls respectively, then calculated proportional reporting ratios (PRR) for the composite of ventricular arrhythmia and cardiac arrest, ventricular arrhythmia only, and QTc-prolongation.

**Results:** Among 18,499,626 unique cases, 61,961 (0.3%) mentioned at least one psychoactive drug of interest. The psychedelic drugs ibogaine, MDA, MDMA, and LSD and the non-psychedelic mitragynine exhibited a significant PRR for ventricular arrhythmia/cardiac arrest; the strongest signals were observed for mitragynine (PRR 8.8; 84/1171 reports), and ibogaine (PRR 38.8; 7/25 reports). When restricting analysis to ventricular arrhythmia alone (excluding potential non-arrhythmic cardiac arrest), the signal remained significant for mitragynine (PRR 4.4, 8/1171 reports) and was markedly higher for ibogaine (PRR 142.0, 5/25 reports), approximately 6-fold higher than dofetilide (PRR 24.0; 533/11,950 reports). This corresponded with QTc-prolongation signals for ibogaine (PRR 121.0; 6/25 reports) and mitragynine (PRR 15.9; 38/1171 reports), which were comparable to dofetilide (PRR 30.9; 771/11,950 reports) and methadone (PRR 12.4; 1428/55,895 reports). MDA, MDA, and LSD did not show ventricular arrhythmia signal, and although MDMA was associated with disproportionate reporting of QTc-prolongation (PRR 9.9; 10/474 reports).

**Conclusions:** Ibogaine and mitragynine demonstrated disproportionate reporting of ventricular arrhythmia similar to or exceeding established proarrhythmic drugs. These data suggest that robust cardiac safety evaluations will be a regulatory priority given accelerated development of psychedelic drugs.

## INTRODUCTION

Illicit drug use has been increasing dramatically throughout the world.(Center for Behavioral Health Statistics, n.d.; European Union Drug Agency, 2025; Ritchie et al., 2022) Simultaneously, there is renewed interest in utilizing certain illicit drugs to treat refractory psychiatric conditions such as treatment-resistant depression and post-traumatic stress disorder (PTSD). These compounds range from plant- and animal-based ancient traditional medicine remedies (e.g., mitragynine, psilocin, ibogaine, mescaline) to modern synthetic compounds like lysergic acid diethylamide (LSD) and 3,4-methylenedioxymethamphetamine (MDMA). Psychedelics are purported to have therapeutic potential as entheogens and psychoplastogens: small molecule drugs that produce rapid and sustained effects on neuronal structure and function and may therefore manifest therapeutic benefit after single dose or intermittent administration.(Volkow et al., 2023)

Although emerging evidence supports potential utility in refractory psychiatric disorders, the US Drug Enforcement Administration (DEA) still categorizes these substances as Schedule I drugs with no accepted medical use and high abuse liability.(US Drug Enforcement Administration, 2026) Some substances have been decriminalized at a local level, resulting in a patchwork of availability and oversight across the US. (**Figure 1**). Mitragynine, a mixed stimulant/opioid, is the principal alkaloid in kratom. Kratom it is not federally scheduled under the Controlled Substances Act, and its status varies by state, ranging from largely unrestricted retail availability -including convenience stores- to prohibition or Schedule I control. None of these substances are approved by the FDA for clinical use at time of this writing, including a recent Complete Response (non-approval) for MDMA.(Center for Drug Evaluation Research, 2024) Despite this federal prohibition, 25 states in the US have considered 74 bills (with 10 enacted) creating an patchwork of expanding utilization across the US.(Siegel et al., 2023) On April 18, 2026, President Donald Trump signed Executive Order 14401 “Accelerating Medical Treatments for Serious Mental Illness,” which has a stated intent to accelerate the development of psychedelic drugs and facilitate access to the treatments for patients with serious mental health conditions.^8^ This Order has potential to provide psychiatric patients with access to potentially beneficial therapies, but aggressive timelines and low rates of certain types of adverse drug events create the potential to miss important safety signals prior to FDA approval.

**FIGURE 1.**
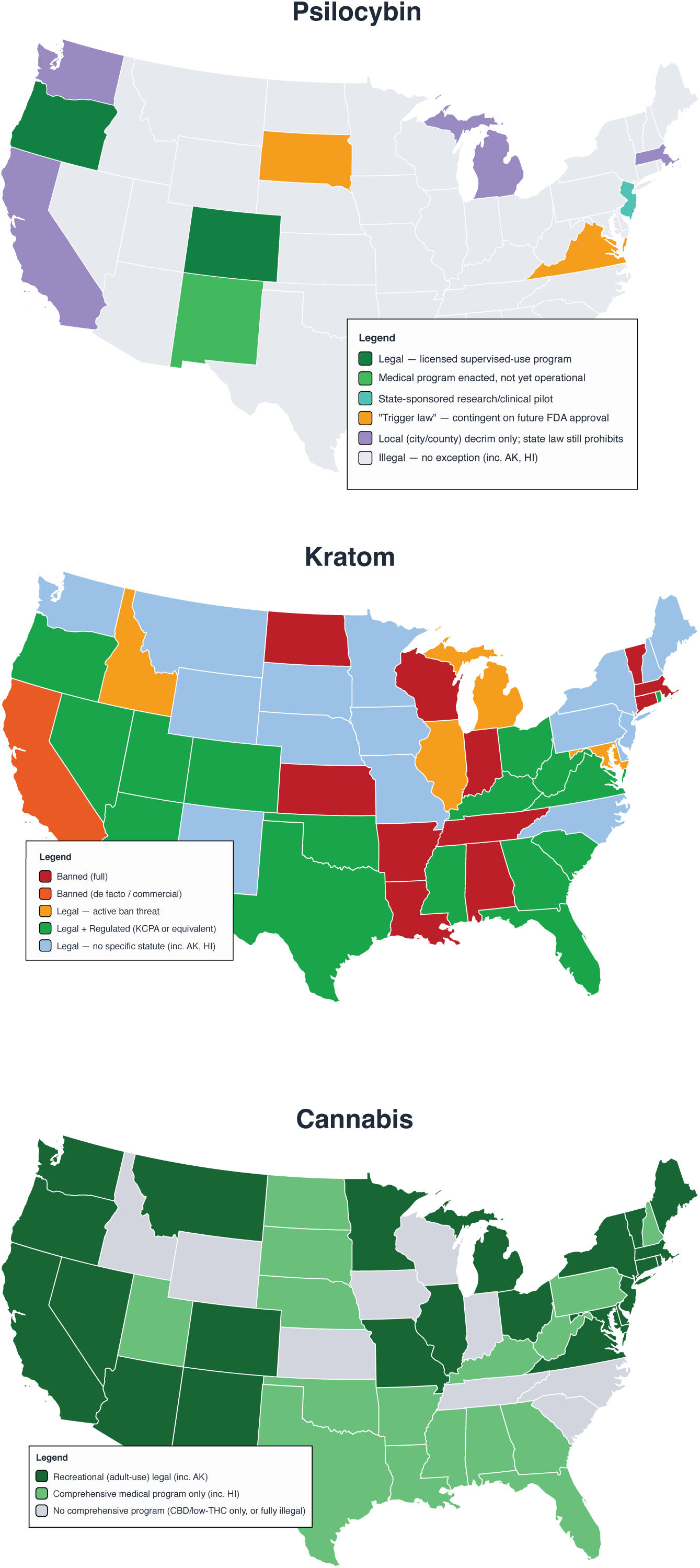
State-level legal regulatory status of psilocybin, kratom, and cannabis, September 2026.

The FDA’s Adverse Event Reporting System (FAERS) is primarily intended to capture adverse drug events (ADEs) from FDA-approved drugs, however, reports also encompass unapproved and illicit substances. Given the lack of systematic evaluation of illicit substances, particularly naturally occurring psychedelics, we assessed whether the known cardiac effects (e.g., sympathetic activation, cardiac ion-channel blockade) might translate into reports of triggered arrhythmia events in FAERS. We analyzed a compendium of Schedule I drugs including those considered solely illicit without accepted therapeutic potential (e.g., cocaine) to psychedelics that have been proposed to have clinical benefit or are being investigated in clinical trials to treat substance use disorders or psychiatric conditions (e.g., MDMA, psilocybin, ibogaine). We investigated whether therapeutically promising Schedule I drugs exhibited disproportionality signals for ventricular arrhythmias, cardiac arrest, and QTc-prolongation.

## METHODS

### IRB approval

All data were fully deidentified by the FDA and are available for unrestricted public download. It was determined that our analysis therefore did not require ethics approval under the National Institutes of Health Human Subjects Research Exemption 4.

### Data source and processing

We obtained publicly available data from FAERS (https://open.fda.gov/data/faers/) through June 2024. FAERS reports include a list of all drugs reported with each adverse event, which may include trade names, combination drugs, and obsolete or foreign names. We matched FAERS cases with active pharmaceutical ingredients using whole and partial string matching as well as limited manual matching in the case of excessive punctuation or special characters and misspelling (DK). Drugs@FDA was used as the reference for FDA-approved drug names and active ingredients as described in FAERS documentation.(*Drugs@FDA Data Files*, n.d.) Ingredient or verbatim drug names were matched to at least one active ingredient from Drugs@FDA in 93.4% (18,937,446 unique cases). Because the drugs of interest in this analysis were not FDA approved, these compounds were identified from verbatim reported drug names using string-matching and manual mapping based on chemical and street names. An additional 5.7% (1,160,557) were therefore matched manually including, but not limited to, the schedule I drugs of interest. Mappings from verbatim text to drugs of interest are provided in the Online Supplement. Unmatched names (e.g. “HC”, “cranberry”, “BLU-U Blue light photodynamic therapy illuminator”) were excluded from this analysis.

To reduce the number of redundant reports, we generated results using unique FAERS case identifiers (*CASEID*) rather than individual FAERS report identifiers (*PRIMARYID*). We analyzed all cases involving schedule I drugs of interest, using all other cases received by FDA during the same time interval for comparison. For example, if the first report involving drug A was received in 2012, cases involving drug A were compared to all other ADE reports starting in 2012, whereas if the first cases involving drug B was in 2010, reports not mentioning drug B starting in 2010 were used for comparison. Adverse events are classified in FAERS using Medical Dictionary for Regulatory Activities (MedDRA) Preferred Terms (PT).(Brajovic, 2010) We analyzed 3 groups of adverse events based on the MedDRA hierarchy (version 25.1) in accordance with previous work:^11,12^ the composite of ventricular arrhythmia/cardiac arrest, ventricular arrhythmia only, and QTc-prolongation. MedDRA PTs comprising each group are enumerated in **Table 1**.

**TABLE 1.** Grouped MedDRA Preferred Terms Analyzed.

| **Ventricular arrhythmia/**<br>**cardiac arrest** | **Ventricular arrhythmias** | **QTc-prolongation** |
| --- | --- | --- |
| Cardiac arrest | Ventricular arrhythmia | ECG QT abnormal |
| Cardiac arrest neonatal | Ventricular tachycardia | ECG QT prolonged |
| Cardio-respiratory arrest | Ventricular tachyarrhythmia | ECG QT interval abnormal |
| Cardio-respiratory arrest neonatal | Ventricular flutter | ECG QTc interval prolonged |
| Parasystole | Ventricular fibrillation | Long QT syndrome |
| Rhythm idioventricular | Torsade de pointes | Torsade de pointes |
| Sudden death |  |  |
| Torsade de pointes |  |  |
| Ventricular arrhythmia |  |  |
| Ventricular asystole |  |  |
| Ventricular extrasystoles |  |  |
| Ventricular fibrillation |  |  |
| Ventricular flutter |  |  |
| Ventricular tachycardia |  |  |
| Accelerated idioventricular<br>rhythm |  |  |
| Sudden cardiac death |  |  |
| Ventricular pre-excitation |  |  |
| Cardiac death |  |  |
| Pulseless electrical activity |  |  |
| Ventricular tachyarrhythmia |  |  |

### Drugs and variables of interest

Schedule I psychedelic drugs of interest were, MDMA (reported in FAERS as midomafetamine), 3,4-methylenedioxyamphetamine (MDA, reported in FAERS as tenfetamine), psilocin, LSD, ibogaine, mescaline, and dimethyltryptamine (DMT). Mitragynine is not a federally scheduled drug but has variable state-specific legal status. THC was recently reclassified as Schedule III at the federal level but was a Schedule I substance throughout the study period. Cocaine, methadone, loperamide and dofetilide were analyzed as positive controls given their known association with QTc-prolongation and torsade de pointes.^11,12^ Negative controls without an established association with arrhythmia were heroin (an opioid) and naltrexone (a nonopioid).

### Confirmatory data analysis

Face-validity of associations between drugs of interest and ventricular arrhythmia in FAERS were determined using the reference “clinical risk of QTc-prolongation” as determined using the CredibleMeds^®^ database.^13^ This dynamic registry is continuously updated wherein drugs with proarrhythmic potential are classified as having “known”, “possible”, “conditional”, risk for ventricular arrhythmia (torsade de pointes). The aforementioned data sources are all clinical or epidemiological. We also evaluated in-vitro electrophysiologic data from the published literarture,^14–19^ specifically, the concentration necessary for 50% inhibition (IC_50_) of the human ether-á-go-go (hERG) potassium channel.

### Statistical analysis

Pharmacovigilance analyses of spontaneously reported adverse event data like those from FAERS generally involve identifying signals of disproportionate reporting (SDR), wherein certain adverse effects are reported more frequently with the drug(s) of interest compared with the reporting frequency of the same adverse effect in all other reports combined. Our primary endpoint was the Proportional Reporting Ratio (PRR), which has been recommended by drug safety monitoring agencies,^20^ and has been validated previously for assessment of drug-induced ventricular arrhythmia.^11,12^. The PRR was calculated as:

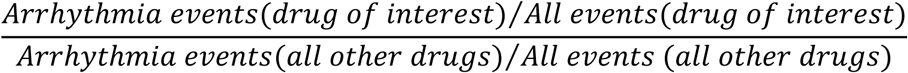

The Empirical Bayes Geometric Mean (EBGM) metric was used as an orthogonal method to reduce likelihood of false positives. The derivation and calculation of the EBGM have been described previously.(DuMouchel, 1999) Briefly, case counts are weighted using a Bayesian shrinkage correction across the entire dataset. This reduces the likelihood of false-positive signals that are due to low numbers of drug-event reports.

Next, we investigated positive and negative controls over the same time periods to facilitate direct comparison with the primary drugs of interest. Data were hosted in Google BigQuery (cloud.google.com, Mountain View, CA). Analyses were performed using RStudio (version 2025.09.02, Posit Software, Boston, MA) and the R statistical package (version 4.4.1, R Foundation for Statistical Computing, Vienna, Austria) through the *bigrquery* package. SDR analyses were performed using the *openEBGM* (version 0.9.1, Canida and Ihrie 2017) and *mdsstat* package (version 0.3.2, ASM Inc., Temecula, CA). FAERS analysis code in R and the BigQuery repository are available on reasonable request. Categorical and continuous variables were compared using the Chi-square (χ^2^) Test and analysis of variance respectively. A p-value < 0.05 was considered statistically significant. The PRR was considered a significant signal of potential regulatory concern if the PRR point estimate was ≥ 2 with χ^2^ statistic ≥ 4, and ≥ 3 reports of the adverse effect of interest associated with the drug. An EBGM was considered significant if the 5% confidence interval was ≥ 2.

## RESULTS

A total of 18,499,626 unique adverse drug event cases were reported from 2000-2024, among which 61,961 (0.3%) mentioned at least one drug of interest. Characteristics of the reported events are summarized in **Table 1**. THC accounted for the the largest number of cases (35,868) whereas ibogaine accounted for the fewest (n=25). PRR and EBGM point estimates for each arrhythmia outcome are summarized in **Figure 2** and **Table 2**.

**FIGURE 2.**
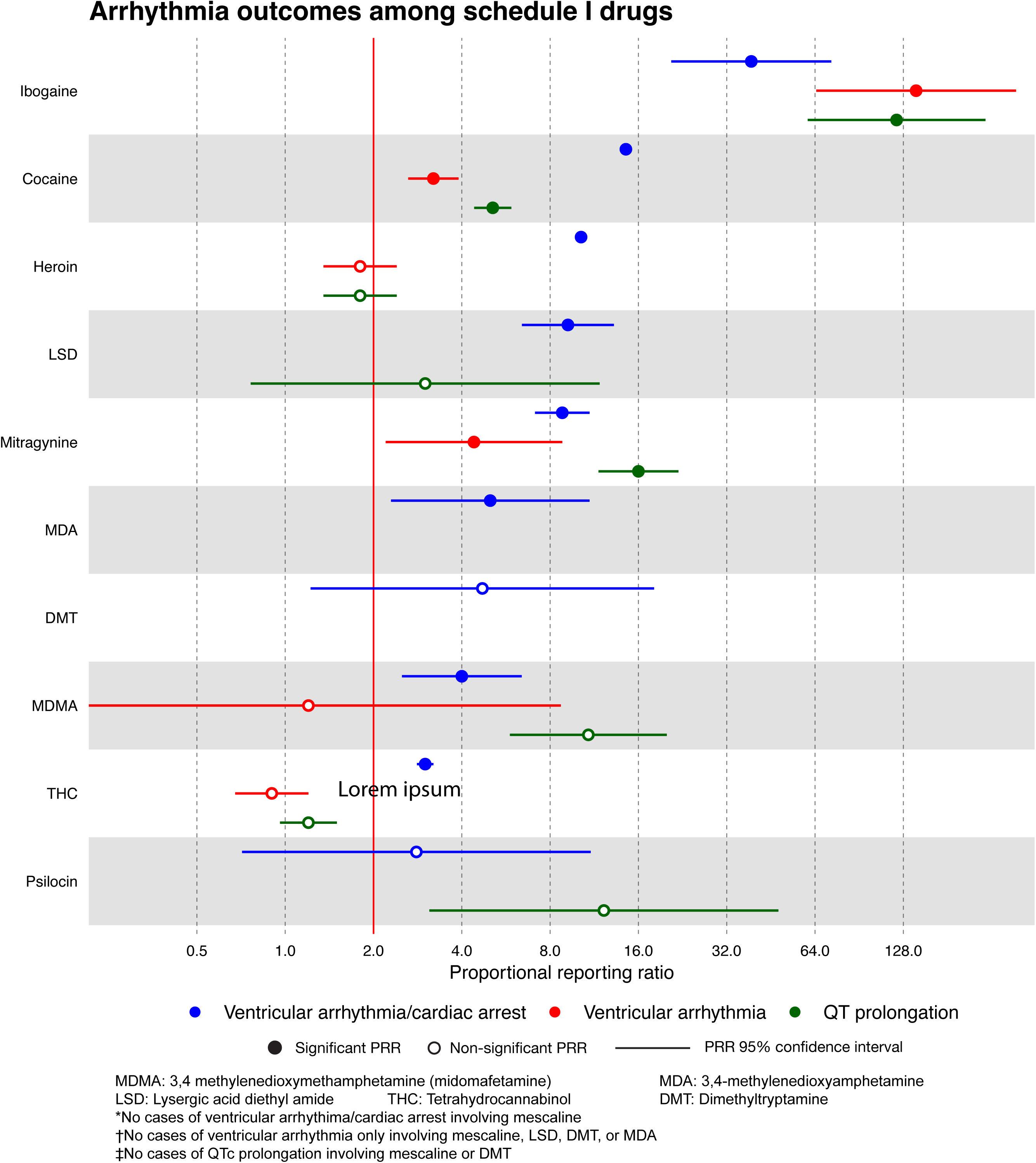
Disproportionate Reporting of Ventricular Arrhythmia and QTc-prolongation Among Psychoactive Drugs

**TABLE 2.** Subject Characteristics, and Arrhythmia Risk Signals for Psychoactive and FDA-Approved Drugs.

| Patient characteristics |  |  |  |  | Ventricular arrhythmia/<br>cardiac arrest |  |  | Ventricular arrhythmia |  |  | QTc-prolongation |  |  |
| --- | --- | --- | --- | --- | --- | --- | --- | --- | --- | --- | --- | --- | --- |
| Drug | Reports<br>(N) | Age, yrs<br>Mean±SD | Male<br>N (%) | Died<br>N (%) | Rate<br>N (%) | PRR | EBGM | Rate<br>N (%) | PRR | EBGM | Rate<br>N (%) | PRR | EBGM |
| Schedule I psychedelic compounds |  |  |  |  |  |  |  |  |  |  |  |  |  |
| Ibogaine | 25 | 32.8±7.9 | 15/24 (62.5) | 20 (80) | 7 (28) | 38.8 | 26.9 | 5 (20) | 142 | 80.8 | 6 (24) | 121 | 89.3 |
| MDA | 135 | 32.6±13.5 | 85/136 (67.5) | 58 (43.0) | 6 (4.2) | 5.0 | NS | 0 (0) | † | † | 0 (0) | † | † |
| MDMA | 435 | 26.5±9.5 | 269/398 (67.6) | 105 (24.1) | 17 (3.6) | 3.9 | 2.41 | 1 (0.2) | † | NS | 10 (2.3) | 10.8 | 4.6 |
| LSD | 317 | 28.1±9.9 | 209/290 (72.1) | 59 (18.6) | 27 (8.3) | 8.9 | 8.7 | 0 (0) | † | † | 3 (0.9) | NS | NS |
| DMT | 48 | 37.0±15.5 | 27/43 (63.6) | 4 (8.3) | 2 (4.2) | † | NS | 0 (0) | † | † | 0 (0) | † | † |
| Mescaline | 40 | 52.4±19.8 | 9/39 (23.1) | 3 (7.5) | 0 (0) | † | † | 0 (0) | † | † | 0 (0) | † | † |
| Psilocin | 78 | 26.6±12.4 | 40/70 (57.1) | 1 (1.3) | 2 (2.5) | † | NS | 0 (0) | † | † | 2 (2.5) | † | 1.1 |
| Schedule I non-psychedelic compounds |  |  |  |  |  |  |  |  |  |  |  |  |  |
| Mitragynine | 1171 | 35.6±11.8 | 788/1049 (75.1) | 429 (36.6) | 84 (7.1) | 8.8 | 6.5 | 8 (0.7) | 4.4 | 3.1<sup>‡</sup> | 38 (3.2) | 15.9 | 15.5 |
| THC | 35868 | 36.9±19.1 | 13212/23628 (55.9) | 3903 (10.9) | 1006 (2.7) | 2.9 | 3.2 | 61 (0.2) | NS | NS | 95 (0.3) | NS | NS |
| Schedule I non-psychedelic positive and negative controls |  |  |  |  |  |  |  |  |  |  |  |  |  |
| Cocaine | 17043 | 36.5±12.5 | 10248/15388 (66.6) | 8920 (52.3) | 2255 (13.2) | 14.5 | 15.0 | 104 (0.6) | 3.2 | 3.3 | 188 (1.1) | 5.1 | 5.3 |
| Heroin | 13586 | 36.3±13.8 | 8324/12013 (69.3) | 7159 (52.7) | 1280 (9.3) | 10.2 | 10.5 | 45 (0.3) | NS | NS | 54 (0.4) | NS | NS |
| FDA-approved controls |  |  |  |  |  |  |  |  |  |  |  |  |  |
| Dofetilide | 11807 | 69.8±11.4 | 5516/10622 (51.9) | 786 (6.7) | 713 (6.0) | 6.5 | 6.9 | 533 (4.5) | 24.0 | 25.4 | 771 (6.5) | 30.9 | 32.4 |
| Methadone | 55529 | 42.7±18.3 | 26403/48029 (55) | 17331 (31.3) | 4246 (7.6) | 8.4 | 8.7 | 827 (1.5) | 8.0 | 8.3 | 1428 (2.6) | 12.4 | 12.2 |
| Loperamide | 78184 | 61.2±17.8 | 29375/72165 (40.7) | 7578 (9.7) | 1695 (2.1) | 2.3 | 2.5 | 839 (1.1) | 5.7 | 5.9 | 1027 (1.3) | 6.2 | 6.4 |

PRR: Proportional reporting ratio
NS: Non-significant – Definitions: PRR > 2 + $\chi^2 > 4 + n \geq 3$
MDMA: 3,4-Methylenedioxymethamphetamine or midomafetamine
LSD: Lysergic acid diethylamide
EB05: 5<sup>th</sup> percentile of Empiric Bayes Geometric Mean
\*Significant EBGM (EB05 ≥ 2)
MDA: 3,4-methylenedioxymphetamine or tenfetamine
THC: Tetrahydrocannabinol
<sup>‡</sup>EB05 = 1.7
† Insufficient cases to estimate
DMT: Dimethyltryptamine

As expected, all positive controls (cocaine, dofetilide, methadone, and loperamide) demonstrated prominent arrhythmia risk signals. Among FDA-approved positive controls, dofetilide exhibited the strongest signal for ventricular arrhythmia alone (PRR=24.0) and QTc prolongation (PRR=30.9). Methadone also demonstrated signals for ventricular arrhythmia alone and QTc prolongation (PRR=8.0 and 12.4, respectively). Loperamide, an FDA-approved over-the-counter drug, showed clear and consistent signals for both outcomes (PRR = 5.7 and 6.2, respectively), representing the lowest magnitude signal among the positive controls (**Table 2**).

Among the drugs of interest, MDMA, LSD, ibogaine, mitragynine, and THC demonstrated significant signals of disproportionate reporting for the broader ventricular arrhythmia/cardiac arrest composite When this outcome was restricted to ventricular arrhythmia alone, however, significant signals persisted only for ibogaine and mitragynine (PRR =142.0 and 4.4 respectively). Both substances also exhibited signals for QTc-prolongation (PRR = 121.0 and 15.9 respectively). The signals for associated with mitragynine were of a magnitude broadly similar to those observed with methadone for ventricular arrhythmia alone (PRR = 4.4 versus 8.0) and QTc-prolongation (PRR = 15.9 versus 12.4). The remaining psychedelic agents (DMT, LSD, mescaline, MDMA, psilocin, MDA) were not significant for either ventricular arrhythmia alone or QTc-prolongation nor was THC.

EBGM results were concordant with the PRR findings. EBGM additionally identified a significant association between MDMA and QTc-prolongation (EBGM=8.7; 90% CI, 4.6–16.1) that was not detected by the PRR methodology. For mitragynine and ventricular arrhythmia alone, the EBGM estimate remained significant but was somewhat attenuated relative to the PRR estimate (EBGM=3.1; 90% CI, 1.7–5.3).

Ibogaine exhibited the highest risk signal of all drugs analyzed for ventricular arrhythmia alone and QTc-prolongation, whereas among FDA-approved drugs, dofetilide was associated with the highest risk signal for ventricular arrhythmia alone (PRR =24.0) and QTc-prolongation (PRR =30.9). Mitragynine and methadone exhibited similar signals for ventricular arrhythmia alone (PRR =4.4 and 8.0, respectively) and QTc-prolongation (15.9 vs. 12.4, respectively). EBGM was concordant with PRR in all cases with the exception of a significant association between MDMA and QTc-prolongation (EBGM 8.7 [90% CI 4.6-16.1]) and a lesser association between mitragynine and ventricular arrhythmia (EBGM 3.1 [90% CI 1.7-5.3]). DMT, LSD, mescaline, MDMA, psilocin, MDA, and THC did not have significant PRRs for either ventricular arrhythmia alone or QTc-prolongation.

Relationships between hERG-blocking potency (IC_50_), clinical classification of QTc-prolongation risk, and PRR values for the adverse events of interest are presented in **Table 3 and Figures 3a-c**, together with positive and negative controls Notably, among the psychedelic drugs with proarrhythmic risk (i.e., ibogaine) and FDA-approved proarrhythmic drugs (i.e., dofetilide, methadone, loperamide), disproportionate reporting rates for ventricular arrhythmias and QTc-prolongation were concordant supporting QTc-prolongation as a plausible mechanism for inducing ventricular arrhythmia.(Woosley et al., n.d.) Agents for which the PRR for ventricular arrhythmia alone exceeded the PRR for the broader ventricular arrhythmia/cardiac arrest composite had lower hERG IC₅₀ values, indicating greater hERG-blocking potency indicative of proarrhythmic potential, and were more likely to have been classified previously as carrying QT-associated risk in the CredibleMeds database.^13^ This pattern was particularly pronounced for ibogaine, for which the PRR for ventricular arrhythmia alone markedly exceeded that for the ventricular arrhythmia/cardiac arrest composite (142.0 versus 38.8). Conversely, agents for which the composite-outcome PRR exceeded the ventricular-arrhythmia-only PRR generally had higher hERG IC₅₀ values representing a greater safety margin, along with an absence of previously described clinical evidence of QT-associated arrhythmia risk.

**FIGURE 3a.**
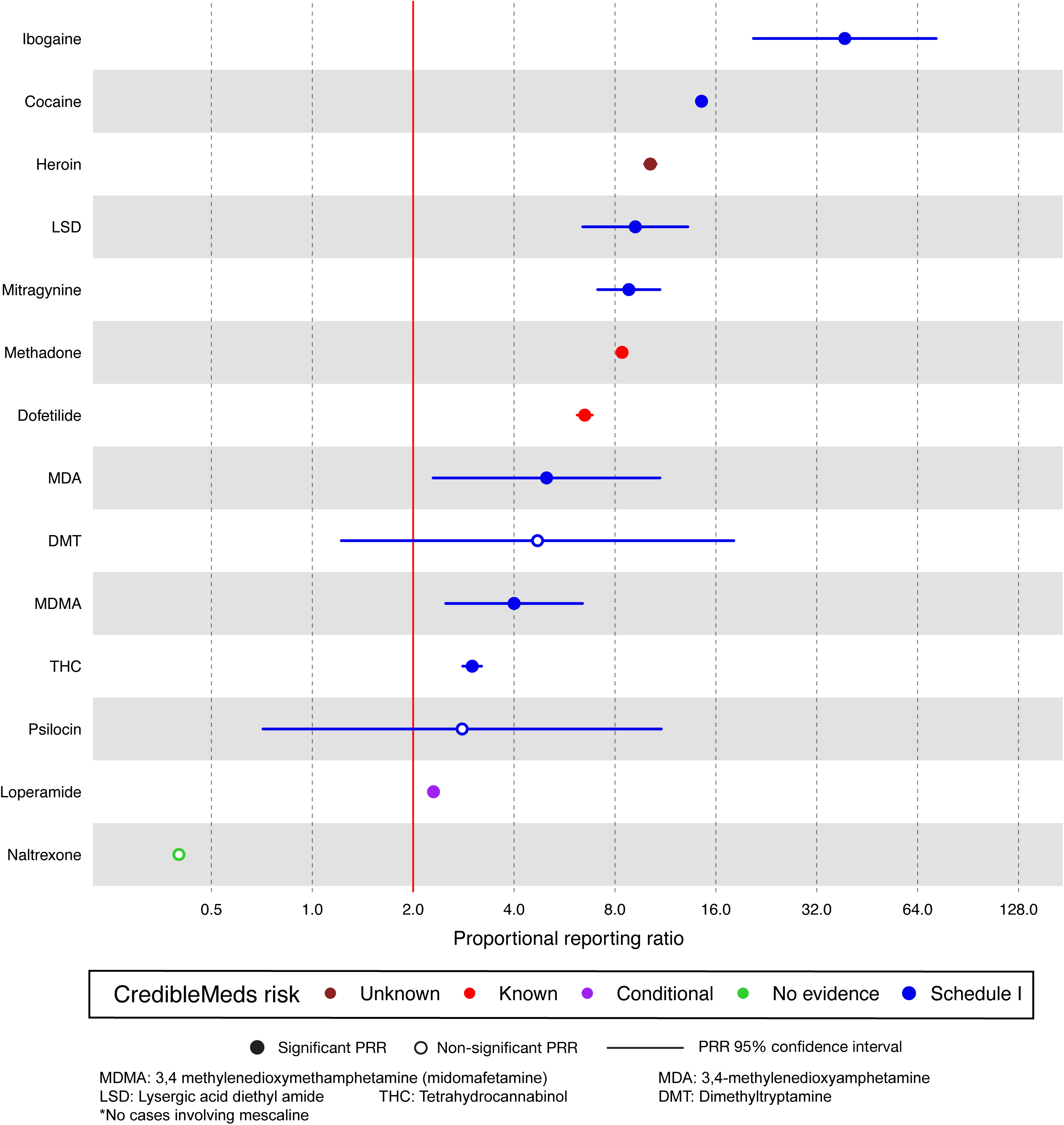
Disproportionate Reporting of Ventricular Arrhythmia/Cardiac Arrest for Psychoactive drugs and Positive and Negative Controls

**FIGURE 3b.**
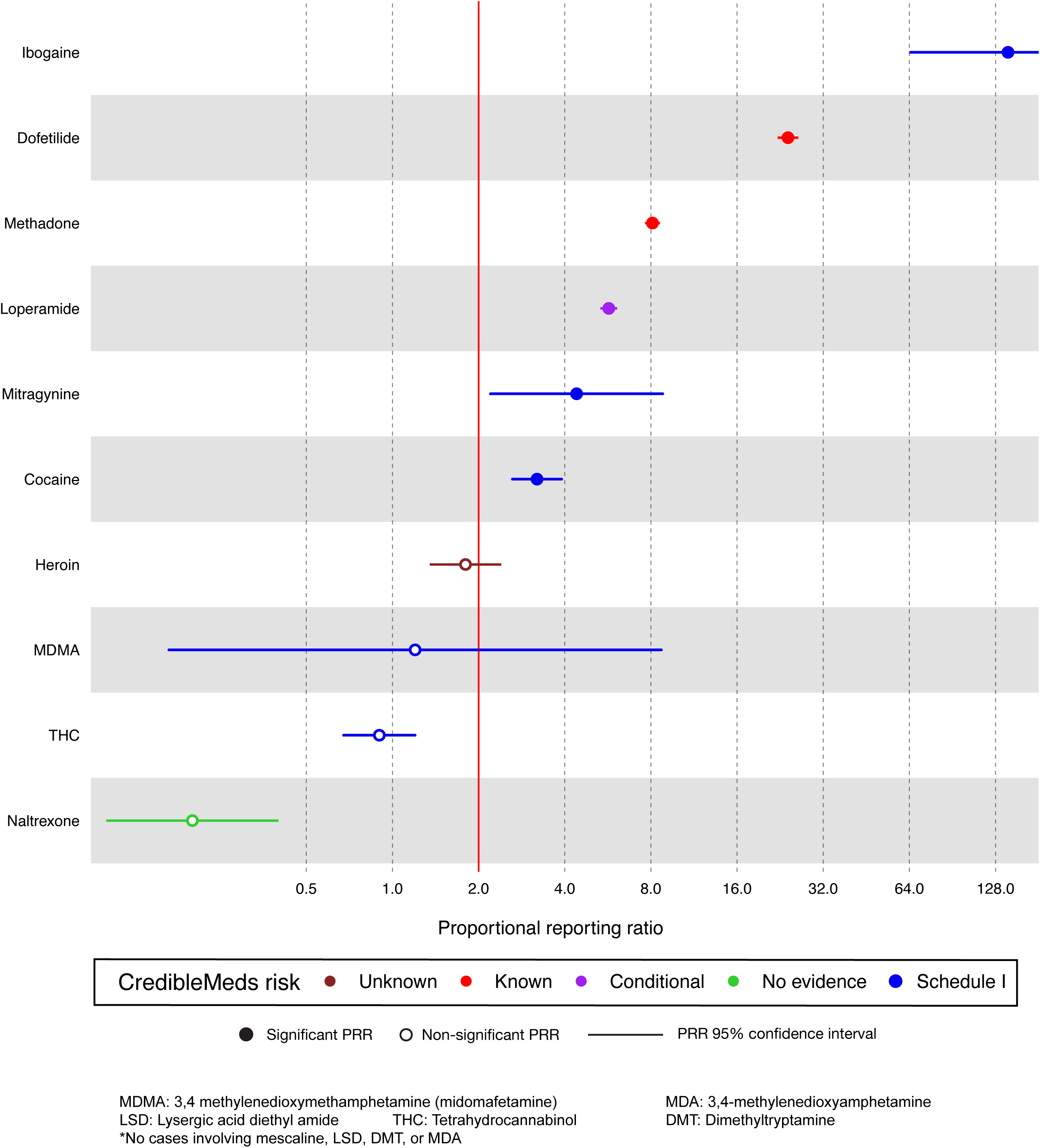
Disproportionate Reporting of Ventricular Arrhythmia Alone For Psychoactive drugs and Positive and Negative Controls.

**FIGURE 3c.**
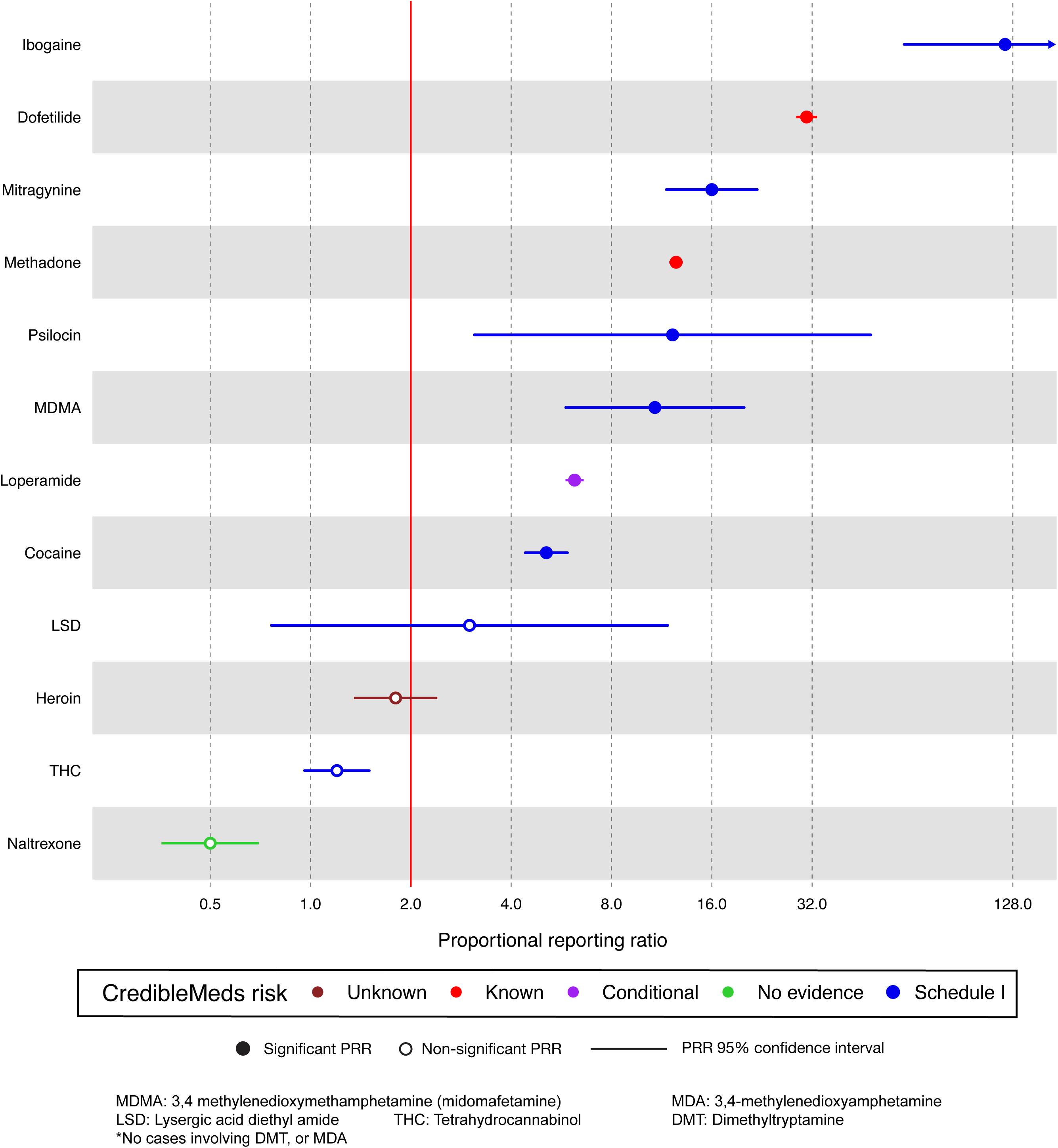
Disproportionate Reporting of QTc-prolongation for Psychoactive Drugs and Positive and Negative controls

**TABLE 3.** Disproportionate Arrythmia Reporting, Clinical Risk of QTc-prolongation, and IC_50_ for hERG Channel Blockade.

| Drug | Arrhythmia Risk* | hERG IC<sub>50</sub> (μM)† | VA | VACA | VA/VACA Ratio |
| --- | --- | --- | --- | --- | --- |
| Ventricular arrhythmia/Cardiac arrest > Ventricular arrhythmia only |  |  |  |  |  |
| Cocaine | Known | 5.60 | 3.2 | 14.4 | 0.2 |
| Heroin | Not yet evaluated | 427 | 1.7 | 10.2 | 0.2 |
| MDMA | Not yet evaluated | 206 | 1.1 | 3.9 | 0.3 |
| THC | Not yet evaluated | 10.3 | 0.9 | 2.9 | 0.3 |
| Mitragynine | Not yet evaluated | 0.90 | 4.4 | 8.8 | 0.5 |
| Ventricular arrhythmia only > Ventricular arrhythmia/Cardiac arrest |  |  |  |  |  |
| Methadone | Known | 4.80 | 8.0 | 8.4 | 1.0 |
| Loperamide | Conditional | 0.09 | 5.7 | 2.3 | 2.5 |
| Ibogaine | Known | 0.50 | 141.5 | 38.8 | 3.6 |
| Dofetilide | Known | 0.04 | 24.0 | 6.5 | 3.7 |

\*From the CredibleMeds® classification of torsadogenic potential are “known,” “possible,” and “conditional,” with regard to the risk of triggering ventricular arrhythmia. †Lower number for IC<sub>50</sub> connotes a higher arrhythmia risk and “lower” safety margin
hERG = human ether a-go-go THC = Tetrahydrocannabinol
MDMA = 3,4-Methylenedioxymethamphetamine (midomafetamine)

## DISCUSSION

There remains a substantial unmet medical need for innovative therapies to treat patients with refractory depression, PTSD, and substance use disorders. However, our pharmacovigilance analysis identified disproportionate signals for proarrhythmia among several psychoactive substances, including DEA Schedule I psychedelic agents under clinical investigation for therapeutic use in these conditions. Signals for ventricular arrhythmia and cardiac arrest were observed with mitragynine (kratom), lysergic acid diethylamide (LSD), 3,4-methylenedioxy-methamphetamine (MDMA), and ibogaine. When the analysis was restricted to ventricular arrhythmia, only mitragynine and ibogaine remained significant. The putative mechanisms underlying proarrhythmia are illustrated conceptually in **Figure 4** and primarily involve 2 mechanisms: sympathomimetic stimulation (among serotonergic agonist drugs such as MDMA and LSD) and hERG-channel blockade (for mitragynine and ibogaine). Unlike ibogaine, however, which promotes polymorphic ventricular arrhythmias solely through QTc-prolongation, mitragynine has also been associated with Brugada-pattern ECG changes and QRS widening, suggesting the possibility of sodium channel blockade or unmasking of latent Brugada syndrome, although a direct effect on cardiac sodium channels has not been established.^11^ Analogously, cocaine exhibited the weakest proarrhythmia signal of all positive controls, which likely reflects the fact that cocaine is a relatively modest hERG-channel blocker—several orders of magnitude less potent than dofetilide—while its predominant myocardial electrophysiologic effect at high concentrations is use-dependent sodium-channel blockade, resulting in QRS widening rather than a manifesting as a QTc-prolongation phenotype.

**FIGURE 4.**
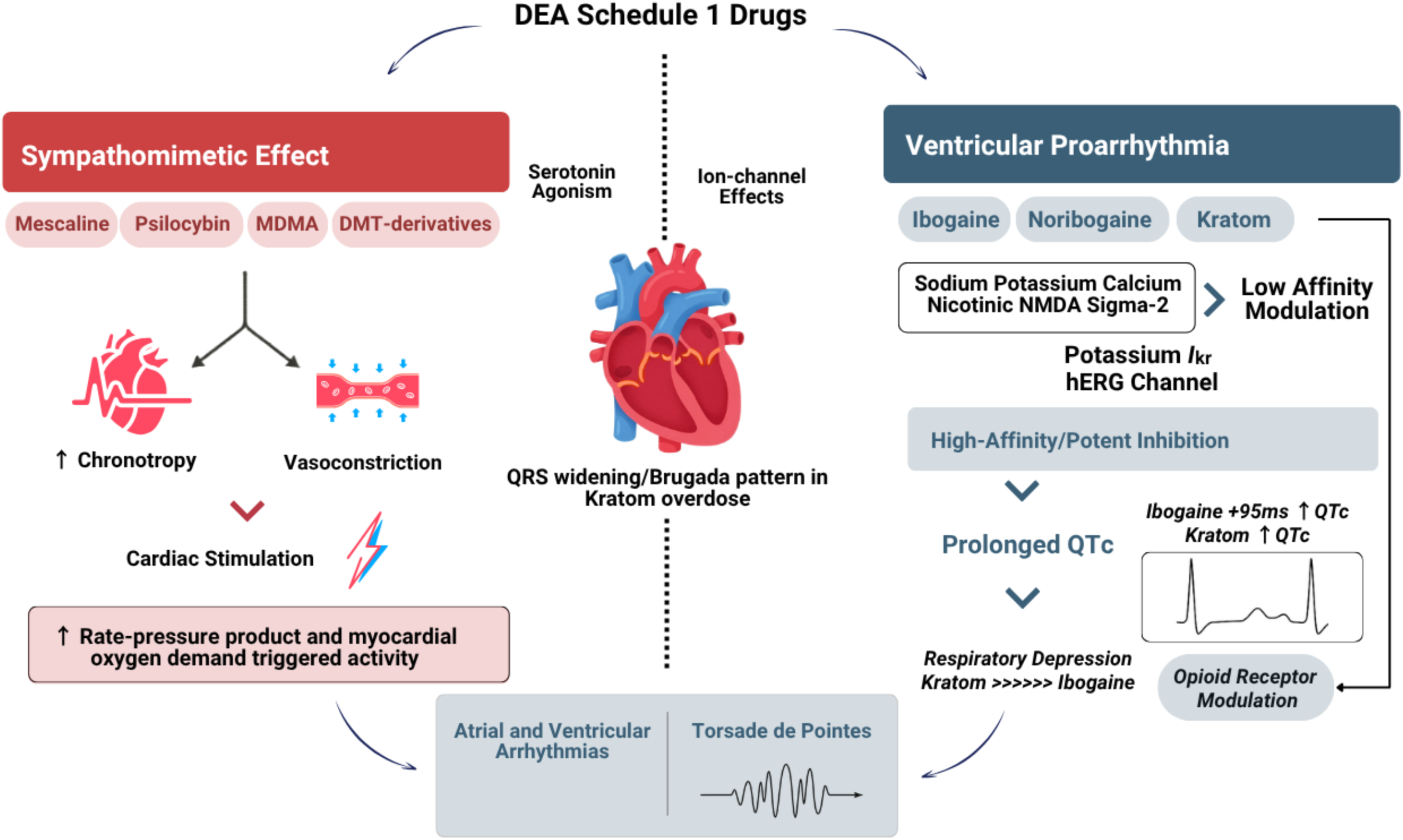
Putative Mechanisms for Arrhythmia Triggering Among Psychoactive Drugs

Several findings from this study warrant consideration from both public health and regulatory perspectives. Among agents with purported therapeutic potential, mitragynine poses a challenging public health threat. This compound exhibited the second strongest proarrhythmic signal in FAERS and is being used chronically without medical oversight by more than 5 million people in the United States.^11^ It is consumed as tea, powder, and capsules and is commonly used to treat acute and chronic pain, opioid withdrawal, anxiety, depression, and PTSD.^21^ These uses persist despite limited evidence of efficacy and sparse safety data. Mitryaginine is extracted from an evergreen tree native to southeast Asia, and ironically is banned in many Asian countries, yet remains widely available in North America through smoke/vape shops, gas stations, convenience stores, and numerous on-line retailers. Its low cost and widespread availability may amplify the associated public health risk, particularly among younger adults; the mean age of patients with arrhythmia events in FAERS was only 35.6 years.

Notably, ibogaine demonstrated the strongest signal for QTc-prolongation and ventricular arrhythmia among all drugs analyzed, particularly after removing the potential confounding influence of unspecified cardiac arrest events. The magnitude of the point estimates for ventricular arrhythmia with ibogaine exceeded those observed for other FDA-approved medications with well-established proarrhythmic potential (i.e., dofetilide and methadone). Current labeling and clinical practice guidelines for these dofetilide and methadone require careful QTc-interval monitoring for safe use,(Ibrahim & Tivakaran, 2025; Krantz et al., 2009) and similar precautions would likely be necessary for ibogaine should it be found to be clinically effective. Ibogaine’s pharmacovigilance signal for arrhythmia was consistent with mechanistic data evaluating cardiac safety margins using ion channel (hERG) assays. This concordance suggests that *in vitro* and rigorous clinical ECG data will be essential to characterize safety during early-phase studies conducted under investigational new drug applications (INDs) in accordance with International Conference on Harmonization best practices,^24^ and prior to conducting larger clinical trials that may support a marketing application. Finally, if ibogaine is ultimately approved, proactive development of educational tools and robust post-marketing surveillance will be essential. This may be particularly important in patients with refractory depression, traumatic brain injury, and PTSD, where concomitant treatment with antidepressants and antipsychotics might further increase QTc-liability.(Cohen et al., 2021; Theal et al., 2020)

The recent Executive Order “Accelerating Medical Treatments for Serious Mental Illness” represents an important opportunity to advance novel treatments for refractory mental health disorders, including PTSD and addiction, among vulnerable populations such as military veterans.(Trump, 2026) Given the ongoing mental health crises affecting these populations, expedited clinical studies and regulatory review could help alleviate substantial suffering. Although intended to apply to any potentially beneficial psychedelic drug, ibogaine is the only specific substance mentioned in the Order, yet our data adds to the observations of others(Grogan et al., 2019; Henstra et al., 2017; Hoelen et al., 2009; Koenig et al., 2013; Meisner et al., 2015; Pleskovic et al., 2012; Steinberg & Deyell, 2018) which suggest that ibogaine is a potent proarrhythmic drug. This is underscored by the fact that disproportionate reporting of ventricular arrhythmia and QTc-prolongation associated with ibogaine was substantially higher than dofetilide, which had a formal risk evaluation and mitigation strategies (REMS) from 2011-2016 years. This included a boxed warning in the prescribing information regarding risk of proarrhythmia, required in-hospital initiation, and implemented a manufacturer-initiated qualified prescriber training program.^22^ Cardiac risks associated with ibogaine might similarly be reduced by via as proactive patient and provider education and induction during continuous telemetry monitoring.

Over the past three decades, there have been a small number of clinical trials investigating the use of psychedelics for challenging or treatment-refractory mental health disorders like substance use disorder, anxiety, depression or PTSD. A number of these candidates are psychedelics, including ibogaine (NCT05029401, NCT04003948, NCT03380728), MDMA (NCT05746572, NCT01458327, NCT05948683), psilocin (NCT05570708, NCT06471959, NCT05065294), LSD (NCT05570708), DMT (NCT06070649), and mescaline (NCT02033707.) Most are small phase 1 or 2 studies with < 50 patients, follow-up ranging from a few hours to several months, and largely rely on symptom surveys or mental health instruments to assess response. Even if these studies progress to phase 3 trials, it is unlikely that those trials will be sufficiently powered for or have long enough follow-up to adequately assess the frequency of rare major adverse cardiac arrhythmia events. If these deficiencies cannot be overcome, it will be important to establish post-approval monitoring plans to ensure adequate patient selection and consistent cardiac safety monitoring during wider implementation.

Because pharmacovigilance is a signal identification tool rather than a definitive scientific assessment, further evaluation was needed to support a potential causal association between psychoactive substances and arrhythmia risk. Accordingly, this study supplemented the FAERS analyses by evaluating in vitro ion-channel and in vivo pharmacokinetic data, comparing hERG IC_50_ values with peak serum concentrations (C_max_). This novel approach found concordance in the observed clinical manifestations relative to the predicted proarrhythmic liability anticipated in vitro. Previous studies have utilized the MedDRA High Level Term (HLT) “Ventricular arrhythmias and cardiac arrest” when assessing arrhythmic potential of drugs in FAERS, as prior evidence has suggested that using standardized groups of terms can improve detection of SDRs at the expense of specificity^12, 34–37^ Compared with those studies that had a significant PRR for “Ventricular arrhythmias and cardiac arrest”, some drugs had an equivalent or larger PRR for ventricular arrhythmia alone or QTc-prolongation (e.g. ibogaine) whereas other substances had smaller or insignificant PRR for ventricular arrhythmia alone or QTc-prolongation despite a high PRR for the composite of ventricular arrhythmia/cardiac arrest. Our finding that compounds with a PRR for ventricular arrhythmia exceeding the PRR for ventricular arrhythmia/cardiac arrest nearly all had lower *in vitro* hERG IC_50_ and were more likely to be classified clinically as increasing risk of QTc-prolongation (CredibleMeds.com).^13^ Conversely, drugs with PRR for ventricular arrhythmia < PRR for ventricular arrhythmia/cardiac arrest had higher hERG IC_50_ and were less likely to have prior clinical evidence of QTc-prolongation effect. We suspected that the HLT “Ventricular Arrhythmias and Cardiac Arrest” may have captured a substantial number of non-arrhythmic events and therefore used manually curated sets of MedDRA Preferred Terms as described herein and suggest this methodology may be an important approach in future studies evaluating proarrhythmic potential

### Limitations

Our study has several limitations common to pharmacovigilance analyses. FAERS is a spontaneous reporting system where only a small fraction of all adverse events that occur are captured, precluding estimation of actual adverse event frequency. Adverse events related to illicit drugs are less likely to be reported and may lack important clinical details. Adverse events related to illicit drugs are also more likely to be severe, given that reporters are often emergency healthcare providers. Illicit drugs often lack standard pharmaceutical or chemical names, and report ascertainment must rely on verbatim street names (e.g., ecstasy for MDMA) from the submitted reports. We included all verbatim drug names mapped to drugs of interest, but it remains likely that some cases were missed. Reports involving illicit drugs are often submitted by patients and providers rather than pharmaceutical companies, creating the possibility that reports tended to be those with more severe outcomes. In addition, age, sex, race, comorbid conditions, anthropometric measurements, and laboratory values were not available in FAERS data and reports are not formally adjudicated by FDA, which may limit accuracy. The two drugs most strongly associated with malignant arrhythmias, mitragynine and ibogaine, have markedly different patterns of use: ibogaine is typically administered as a single dose, whereas mitragynine is often consumed chronically, including multiple times daily. These differences introduce additional uncertainty in characterizing their real-world risks and identifying optimal risk-mitigation strategies. Despite these limitations, we contend that the complementary use of orthogonal pharmacovigilance approaches, ion-channel data, and a widely accepted, dynamic clinical registry of QTc liability strengthens the validity of our findings.

In conclusion, analysis of FAERS data identified disproportionate reporting of ventricular arrhythmias and/or QTc-prolongation with certain psychoactive drugs, including schedule 1 psychedelic drugs, particularly those derived from plant substances, such as ibogaine, and mitragynine (kratom). As efforts to establish the therapeutic efficacy of psychedelic agents continue to expand, equally rigorous evaluation of their cardiovascular safety will be essential to define their benefit–risk profile.

## Data Availability

All data used in this analysis are available for public download from the FDA Adverse Event Monitoring System (FAEMS), formerly known as the FDA Adverse Event Reporting System (FAERS).

https://fis.fda.gov/extensions/FPD-QDE-FAERS/FPD-QDE-FAERS.html

## Acknowledgements

The authors thank Teresa Buracchio of the Center for Drug Evaluation and Research and the Office of Cardiology, Hematology, Endocrinology, and Nephrology leadership for their careful review and clearance of the manuscript.

